# Usefulness of autofluorescence detection of the parathyroid glands with PTeye™ system. Scope review

**DOI:** 10.1101/2025.02.01.25321520

**Authors:** José Luis Pardal-Refoyo, Beatriz Pardal-Peláez

## Abstract

**Introduction and Objective:** Near-infrared (NIR) fluorescence is used to visualize anatomical structures and physiological activities in real time. The objective is to synthesize the advantages and disadvantages of the PTeye™ system in the identification of parathyroid glands during thyroid and parathyroid surgery.

**Method:** A literature review of the scope of primary research articles in databases such as PubMed, WoS and Cochrane was carried out, using the search strategy “(autofluorescence AND (parathyroid glands)) AND PTeye™” until January 27, 2025. We included studies with any methodology that evaluated the identification of parathyroid glands with autofluorescence in thyroid or parathyroid surgery with PTeye™ technology. Review articles, editorials, and individual clinical cases were excluded.

**Results:** We included 8 studies that met the inclusion criteria. The results showed that PTeye™ has high accuracy in identifying parathyroid glands, with identification rates above 90%. The use of PTeye™ reduced the need for additional tests such as freeze biopsies and increased the surgeon’s confidence in identifying parathyroid tissue. However, some false positives were identified, including thyroid nodules and lymph nodes. The device proved to be easy to use and provided real-time feedback.

**Conclusions:** PTeye™ is a useful tool to improve the identification of parathyroid glands during surgery, increasing surgeon confidence and reducing the rate of postoperative hypocalcemia. Although it has some disadvantages, such as the possibility of false positives and the need for a learning curve, its benefits outweigh these limitations. More studies are needed to confirm these findings and assess their long-term impact.

## Introduction

Near-infrared (NIR) fluorescence occurs when fluorophores absorb light at a specific wavelength, emit light at a longer wavelength, and this is detected after interacting with biological tissue [1].

This process consists of several steps [1]. First, during excitation, the NIR fluorophore is activated by a light source, usually a laser at a wavelength specific to each fluorophore (e.g., indocyanine green with 780-810 nm, methylene blue with 660-680 nm, IRDye800CW with 774-789 nm, quantum dots with 700-900 nm, and rare earth nanoparticles with 808-980 nm light). The excitation light is absorbed by the fluorophore, elevating it to an excited energetic state. Then, in the emission phase, after a period in the excited state, the fluorophore returns to its ground state, releasing the absorbed energy in the form of light. This emitted light has a longer wavelength than excitation light and, in the case of NIR fluorophores, is in the near-infrared range (700-1700 nm). Subsequently, both the excitation light and the emitted light must pass through the biological tissue, where they can be reflected, scattered, absorbed, or interfered with by tissue autofluorescence. The NIR window is beneficial due to the lower scattering and absorption of light in this range, allowing for greater tissue penetration and a better signal-to-noise ratio. Finally, the light emitted by the fluorophore is captured by a detector, such as a NIR-sensitive camera, which converts the light into an image or signal (visual or acoustic). The image can be used to visualize anatomical structures, specific biomarkers, and physiological activities in real time.

Several NIR fluorophores are used in biomedical imaging, including organic fluorophores, SWCNTs (single-walled carbon nanotubes), QDs (quantum dots), RENPs (rare earth nanoparticles), and IFPs (fluorescent infrared proteins). In addition, metallic nanoclusters such as gold, polymer nanoparticles conjugated with emission in the NIR and nanoparticles are used for in vivo cell monitoring [1].

There are two near-infrared (NIR) fluorescence emission windows used in biomedical imaging: NIR-I and NIR-II [1].

NIR-I (700-900 nm): This spectral region is traditionally known as the “near-infrared biological transparency window” due to low absorption by tissues and minimal background autofluorescence compared to the visible spectrum. Fluorophores in the NIR-I region include indocyanine green and methylene blue, both of which have been approved by the FDA for clinical use. These fluorophores are used in structural visualization of anatomical features, functional imaging of cardiac perfusion, or for intraoperative surgical guidance [1].

NIR-II (1000-1700 nm): This window reduces tissue dispersion, absorption, and autofluorescence. NIR-II fluorophores include Ag2S quantum dots, rare earth-doped nanoparticles, and single-walled carbon nanotubes (SWCNTs), and are used for deep anatomical, molecular, and functional imaging. These windows allow greater penetration into tissues and better image resolution, being valuable for biomedical research and clinical applications. In addition, optical subwindows such as NIR-IIa (1300-1400 nm) and NIR-IIb (1500-1700 nm) offer additional improvements in clarity and signal-to-noise ratio [1].

The use of NIR fluorescence has several applications [1]. Improved contrast in anatomical, molecular, and functional images allows for better visualization of tumors, identification of biomarkers, and evaluation of physiological activities for diagnosis and during surgery.

To improve the visualization of the parathyroid glands, several external fluorophores have been used in NIR fluorescence imaging, such as indocyanine green (ICG) is excited by light in the range of 780-810 nm and fluoresces around 822 nm; methylene blue (MB) is excited by light in the range of 660-680 nm and fluoresces around 686 nm; the IRDye800CW which is excited by light in the range of 774-789 nm and fluoresces around 789 nm; quantum dots (QDs) that can be excited with light in the range of 700-900 nm and fluoresce in the NIR range; Rare Earth nanoparticles (RENPs) that are typically excited by light in the range of 808-980 nm and fluoresce in the range of 1,000-1,600 nm, all approved by the FDA and used in high-resolution imaging of anatomical and vascular structures [1].

These external fluorophores significantly improve the contrast and clarity of images of the parathyroid glands, making it easier to identify and differentiate them from other tissues during surgical and diagnostic procedures [1].

On the other hand, tissues contain natural fluorophores that emit light when excited and have various biological roles [1]. Some examples are chlorophyll in plants, responsible for photosynthesis and emitting red fluorescence; flavins in enzymes and cofactors such as FAD and FMN, which fluoresce blue to green; NADH, a cofactor in cellular reactions, which emits blue fluorescence; fluorescent proteins such as GFP, used in research to mark cell structures; and bile pigments such as bilirubin, which fluoresce yellow to green.

Both the detection of natural autofluorescence and that emitted by external fluorophores has increased the accuracy in tissue identification. The development of NIR-II fluorophores increases accuracy by improving the ratio of signal (the one emitted by the excitation of the external fluorophore) to the noise (the fluorescence emitted by natural fluorophores) [1].

Natural fluorophores are useful in various biomedical imaging applications and cell biology studies due to their ability to emit light in a targeted and detectable manner [1].

To avoid interference of autofluorescence with external fluorophores in fluorescence imaging, the use of fluorophores that emit in the near-infrared (NIR) is recommended, especially in the NIR-II window (1000-1700 nm), due to their lower dispersion, absorption, and autofluorescence of tissues.

The parathyroid glands have intense autofluorescence under NIR light, significantly greater than that of surrounding tissues, such as the thyroid gland and adipose tissue [1]. This allows the glands to be identified during thyroid and parathyroid surgery. Autofluorescence is due to natural fluorophores such as riboflavin (vitamin B2) and NADH (reduced nicotinamide adenine dinucleotide), known to fluoresce when excited by light in the near-infrared (NIR) range [1].

Riboflavin and NADH fluoresce in the ranges of 500-600 nm and 460 nm respectively, outside the NIR spectrum (700-1700 nm), when excited by light of 450-500 nm and 340-360 nm [1].

The detection of natural autofluorescence is advantageous as it does not require external agents, reduces costs and time, facilitates real-time evaluation, minimizes interference, and is useful in both research and clinical contexts.

Identification of the parathyroid glands by NIR autofluorescence during total thyroidectomy increases the identification rate and reduces the incidence of transient and global postoperative hypocalcemia [2].

Systems are marketed using cameras of different types and sensitivities [3] for imaging (such as Fluobeam® 800, Elevision® IR and PDE Neo II)® or probes for direct tissue contact and detecting the autofluorescence signal without imaging (such as PTeye™®) [2].

Direct contact systems such as PTeye™ have the advantage of being easy to handle during surgery.

The aim of this review is to provide a synthesis of the advantages and disadvantages of the PTeye™® contact system for the detection of the parathyroid glands during thyroid and parathyroid surgery.

## Methods

Bibliographic review of the scope of primary research articles in the PubMed, WoS and Cochrane databases with the search strategy *“(autofluorescence AND (parathyroid glands)) AND PTeye™”* published up to January 27, 2025.

Inclusion criteria: Primary research studies with any methodology (retrospective, prospective, clinical trial) including identification of the parathyroid glands with autofluorescence in thyroid surgery or parathyroid with PTeye™® technology.

The PRISMA-ScR (Preferred Reporting Items for Systematic reviews and Meta-Analyses extension for Scoping Reviews) guidelines were followed for the study (https://www.prisma-statement.org/scoping).

Exclusion criteria: review articles, editorials, individual clinical cases.

The method, the variables studied and the results were evaluated in the articles.

A narrative synthesis was made based on the selected articles.

NotebookLM (https://notebooklm.google.com) was used for text analysis and synthesis table generation.

The level of evidence and degree of recommendation were evaluated with the GRADE tool (https://www.gradepro.org/).

## Results

After initial search and deletion of repeat records, we included 8 studies that met the inclusion criteria [4– 11] as summarized in the PRISMA diagram in Figure 1.

**Figure 1.**
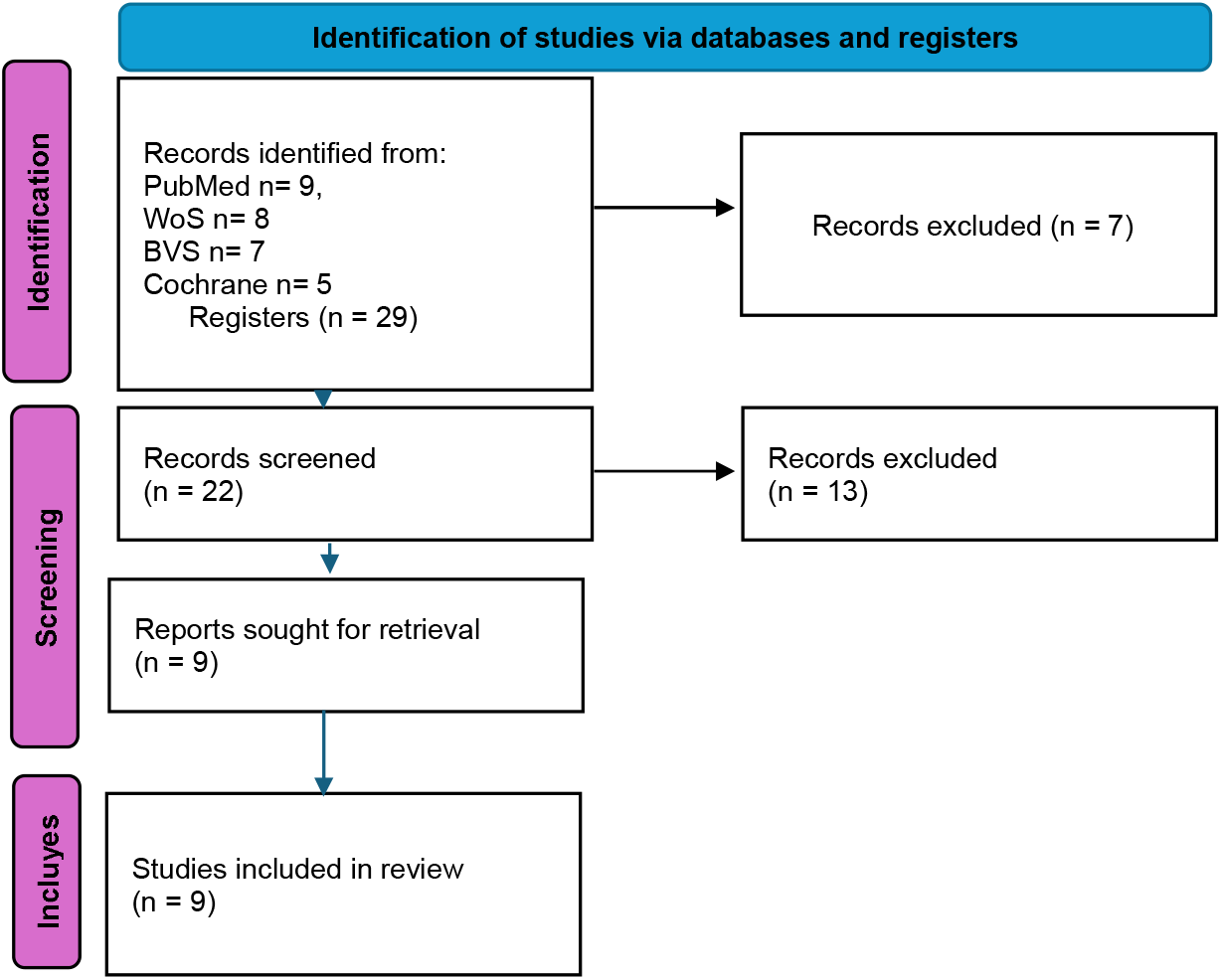
PRISMA diagram with the selection of articles.

Table 1. (Annex 1) summarizes the key characteristics of the selected articles (method, variables studied and main results).

**Table 1.**
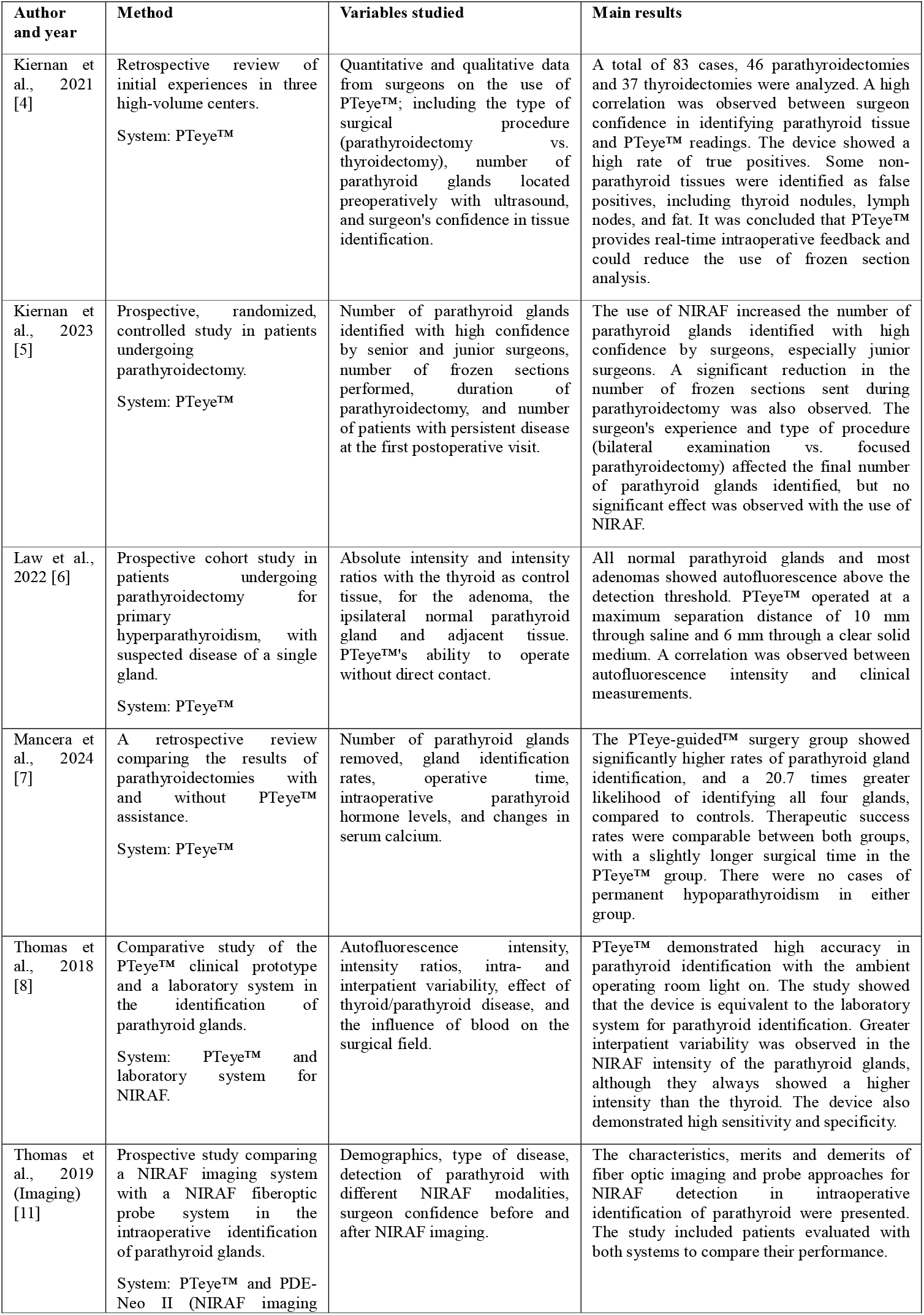
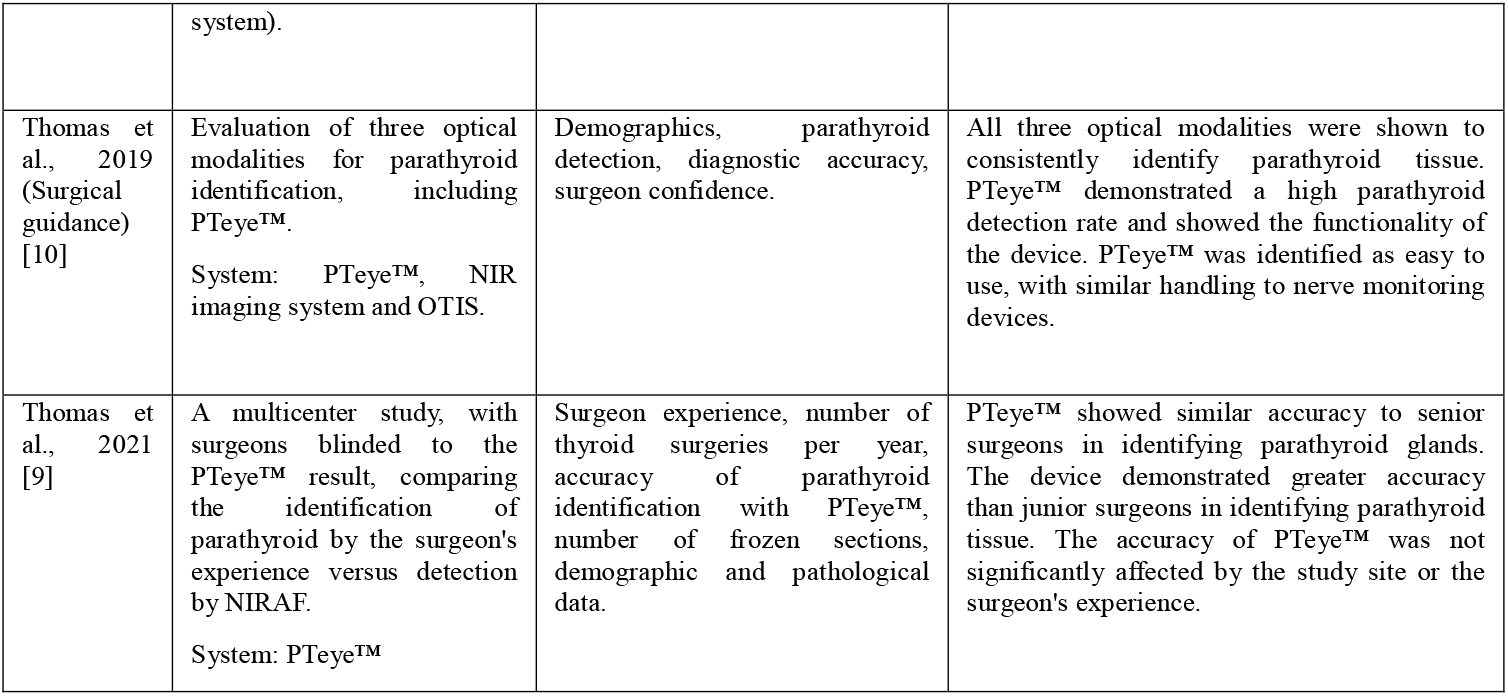
Summary of the results in each study.

## Discussion

PTeye™ employs a fiber-optic probe that is placed in direct contact with the tissue[4,6,10], emits near-infrared light and measures tissue autofluorescence, providing the surgeon with a detection ratio (DR) reading, a RD greater than 1.2 indicates that the tissue is likely parathyroid [6]. The device also includes a console with a display and a foot pedal to activate the measurement [4].

The studies have different methodologies that prevent comparisons to be made about their results. They describe the development and evaluation of prototypes for intraoperative identification of parathyroid glands using near-infrared autofluorescence (NIRAF): PTeye™, a high-precision portable device that works by contact with ambient light; a modified near-infrared imaging system (NIRIS) that provides spatial imaging and an overlay tissue imaging system (OTIS) that projects images directly onto the surgical field.

The results show high accuracy in identifying the parathyroid glands with each system, reducing their accidental removal and postoperative hypocalcemia.

Each device has advantages and disadvantages in terms of ease of use, spatial information, and resistance to light interference.

- PTeye™ [4–11]: It is a probe-based detection device that uses near-infrared autofluorescence (NIRAF) to identify parathyroid tissue. It consists of a console, a sterilizable fiber optic probe, and a foot pedal to activate the measurement. It provides real-time information about the autofluorescence intensity of the tissue and the likelihood that it is parathyroid.
- Laboratory system: In the study by Thomas et al. (2018) [8], a laboratory system including a spectrometer, a 785 nm laser, a custom fiber optic probe, and a computer is used, which was the starting point for the development of PTeye™.
- PDE-Neo II [11]: An imaging system that uses a camera to detect near-infrared autofluorescence and display images on a monitor. Unlike PTeye™, it is not a contact system.
- OTIS (Overlay Tissue Imaging System)[10]: It is an imaging system that projects a visible image directly onto the surgical field, using autofluorescence in the near infrared.

Kiernan (2021) and Thomas (2021) highlight the high accuracy of the PTeye™ device in the identification of parathyroid glands, with concordances of 94.1% and 92.7%, compared to visual identification [4,9].

The accuracy of the PTeye™ system prototype was 96.1% compared to a laboratory system, highlighting its ease of use under operating room lights [8]. In addition, the device improves confidence in identification without direct contact and suggested autofluorescence differences between adenomas and normal glands [5,6], with higher identification rates, no incidence of permanent hypoparathyroidism, and its usefulness in identifying lower glands [7]. PTeye™ offers greater sensitivity and accuracy than PDE-Neo II and is useful for locating unidentified glands preoperatively[11].

These results suggest that PTeye™ is a useful and accurate tool for intraoperative identification of parathyroid glands that offers advantages and disadvantages (Table 2).

**Table 2.** Advantages and disadvantages of the PTeye™ autofluorescence device.

| Advantages of PTeye™ | Drawbacks of PTeye™ |
| --- | --- |
| Real-time identification | False positives (cancerous thyroid tissue, thyroid nodules, lymph nodes, and brown fat) |
| Reduces the need for intraoperative testing (freeze biopsies for confirmation of parathyroid tissue or analysis of PTH levels) | Learning Curve |
| Increases surgeon confidence | Dependence on the surgeon's skill |
| Educational tool |  |
| Save time in the operating room | Limitations in secondary hyperparathyroidism |
| High accuracy in the identification of the parathyroid glands (greater than 90% with high positive and negative predictive values). | Need for preview viewing |
| Works in the presence of ambient light | Dense blood clots may block the signal. |
| Usefulness in various surgeries: It is beneficial in reoperation surgeries, cases of non-localized parathyroid, and in patients with Hashimoto's thyroiditis. It is also useful in identifying at least one residual PG after a total thyroidectomy. |  |
| Reduces postoperative morbidity |  |
| Works without direct contact: The device works properly even if the probe is not in direct contact with the tissue, up to 10 mm through saline and 6 mm through clear solid medium. |  |
| Benefits in the location of the lower glands that have a more variable location. |  |
| May improve gland identification in patients with multiglandular disease |  |

PTeye™ offers significant advantages in parathyroid and thyroid surgery, primarily due to its ability to provide real-time identification of parathyroid tissue [8], allowing surgeons to make more informed intraoperative decisions. This immediate feedback decreases the need for additional tests such as freeze biopsies (FSA) or aspiration to measure PTH [4]. The use of PTeye™ also increases the surgeon’s confidence, regardless of their level of experience, confirming parathyroid tissue even in cases of high initial confidence [5]. In addition, it works as a real-time educational tool, saving time in the operating room by avoiding additional procedures and facilitating the identification of the parathyroid glands [4]. PTeye™’s accuracy is remarkable, with studies demonstrating greater than 90% sensitivity and specificity [6]. Its ability to operate in ambient light, unlike some imaging systems, and without direct tissue contact, to some degree, also represents a practical advantage [11] [10]. In addition, it facilitates the location of the lower glands and the identification of the four glands, important in patients with multiglandular disease [7]. It is also useful in reoperations, cases of non-localized parathyroid, and in patients with Hashimoto’s thyroiditis [4].

However, PTeye™ also has some disadvantages. It can generate false positives with cancerous thyroid tissue, thyroid nodules, lymph nodes, and brown fat, requiring careful interpretation of the results [4]. There is a learning curve to its effective use, and the need to establish an appropriate baseline is necessary to obtain reliable results [4]. The results may be misinterpreted by less experienced personnel, or trainees may become overly dependent on the device rather than their own surgical judgment [4]. The device may be less effective in patients with secondary renal disease-induced hyperparathyroidism [8]. In addition, it requires the surgeon to first visualize the suspicious tissue before confirmation, as it does not provide spatial information of the area [10,11]. Although some studies suggest that blood does not affect the NIRAF measurement, bleeding or dense clots could obstruct the signal. Interpretation of results should be combined with surgical experience and other diagnostic tests, such as freeze biopsy and analysis of intraoperative PTH levels [8].

### Implications for the GRADE assessment

Level of evidence: The evidence is moderate to high due to the presence of prospective and comparative studies, although some are retrospective. The consistency of the results strengthens the level of evidence. The presence of some studies with control groups also contributes to a higher level of evidence. However, the non-randomised nature of some studies slightly decreases the level of evidence

#### Recommendation

A recommendation in favor of the use of PTeye™ in thyroid and parathyroid surgeries is likely to be considered, given the demonstrated benefits in identifying glands, improving surgeon confidence, and usefulness in training. Proper training and correct device configuration are necessary to minimize false positives and negatives, especially in cases of adenomas and patients with secondary hyperparathyroidism. The recommendation would likely be conditional in favour due to methodological limitations in some studies and the need for more research on long-term outcomes in patients.

Additional research needs:

Evaluation of the long-term impact on patient outcomes (e.g. postoperative hypocalcemia, success of parathyroidectomy).

Randomised controlled studies to further strengthen the level of evidence.

Research on the use of PTeye™ in different types of parathyroid pathologies and different populations.

Resource use impact assessment (e.g. reduction of biopsies by freezing).

Cost-effectiveness analysis of PTeye™.

In summary, although a complete GRADE assessment cannot be performed with the information provided, studies suggest that PTeye™ is a tool that improves intraoperative identification of the parathyroid glands, with a moderate to high level of evidence and a conditional recommendation in favor of its use. The specific clinical context needs to be considered and further research needs to be conducted to confirm these findings.

### Limitations of the bibliographic review on the selected studies

#### Sample size and generalizability

Some studies have a limited sample size, which restricts generalizability of results to broader populations [4–6]. For example, some studies were conducted at a single center or with a small number of surgeons, which may not be representative of the diversity of surgical practices [4,6]. In addition, the exclusion of patients with secondary hyperparathyroidism in some studies may affect the applicability of the results [10],

#### Lack of standardization in procedures

Variability in surgical approaches among surgeons can complicate the interpretation of results. For example, bilateral neck exploration (BNE) compared to selective parathyroidectomy may influence the number of glands identified [7]. The lack of standardization in procedures is also related to different surgical techniques, which can make it difficult to compare results between studies [4].

#### Confounding variables

The operative time in parathyroidectomy can be affected by multiple factors, such as previous radiological location, the anatomy of the gland, the presence of ectopic or supernumerary glands, the size of the gland, and the surgeon’s experience. This makes it difficult to determine the actual effect of probe-based NIRAF detection on operative time [7].

#### Technology limitations

Penetration of NIR wavelengths is limited, making it difficult to detect parathyroid glands located deep in or under layers of fatty or thyroid tissue [6]. PTeye™ technology does not assess parathyroid perfusion/viability, only its presence [4]. PTeye™ results may be influenced by elevated reference settings, and the adenomatous region of a gland may show low indices due to the heterogeneity of NIRAF [4]. Interpreting the results of new technologies like PTeye™ requires surgical expertise, and trainees can accept false positive results, relying more on the device than on their own experience [4].

#### Surgeon bias

Interpretation of results can be subjective and error-prone, and surgeons, especially trainees, may rely more on the device than on their own surgical judgment [4]. In some studies, surgeons were not blinded to the use of NIRAF technology, which could bias their interpretation of the results[5].

Lack of long-term follow-up: Some studies lack data on long-term surgical success and failure, as only a portion of the cohort had a follow-up of 6 months or more [4].

#### Selection bias

Studies may have a selection bias because patients with radiologically localized disease were considered for selective parathyroidectomy, while non-localized cases underwent bilateral neck examination [4].

#### Limitations in validation

Histology served as validation only for excised specimens, without a gold standard for validating healthy parathyroids that are left *in situ*. In addition, tissues identified with low confidence without corresponding histology were excluded from the analysis [4,9].

#### Lack of cost-effectiveness analysis

Although it has been suggested that the use of technologies such as PTeye™ can reduce costs by minimizing the need for freeze biopsies, further cost-benefit evaluation is needed in various settings [4].

#### Interpatient and intrapatient variability

There is variability in the intensity of NIRAF between patients and even within the same patient. Thyroid or parathyroid disease may influence parathyroid autofluorescence [6].

#### Limitations in tissue detection

The presence of blood in the surgical field may affect the measurement of NIRAF. It was also observed that thyroid tissue tends to have a higher NIRAF intensity than fat, which could mask the NIRAF intensity of an underlying parathyroid [4] [4,6,10].

These limitations need to be considered when evaluating results and planning future research, as NIRAF technology such as PTeye™ complements but does not replace the surgeon’s expertise [4].

## Conclusions

PTeye™ is a useful tool to improve the identification of parathyroid glands during surgery, both primary and in reinterventions, which increases surgeon confidence and can reduce the rate of postoperative hypocalcemia.

It is a simple operating system that works by contact with ambient light.

It can reduce the need for freeze biopsies and thus save time and costs in surgery. Specific cost studies are lacking.

More studies with a higher level of evidence are needed to demonstrate a significant impact on clinical outcomes.

## Data Availability

All data produced are available online at

## ANNEX 1

